# Executive function deficits in genetic frontotemporal dementia: results from the GENFI study

**DOI:** 10.1101/2024.05.16.24307390

**Authors:** Lucy L Russell, Arabella Bouzigues, Rhian S Convery, Phoebe Foster, Eve Ferry-Bolder, David M. Cash, John C. van Swieten, Lize C. Jiskoot, Harro Seelaar, Fermin Moreno, Raquel Sanchez-Valle, Robert Laforce, Caroline Graff, Mario Masellis, Maria Carmela Tartaglia, James B. Rowe, Barbara Borroni, Elizabeth Finger, Matthis Synofzik, Daniela Galimberti, Rik Vandenberghe, Alexandre de Mendonça, Chris Butler, Alexander Gerhard, Simon Ducharme, Isabelle Le Ber, Isabel Santana, Florence Pasquier, Johannes Levin, Sandro Sorbi, Markus Otto, Jonathan D. Rohrer, the Genetic FTD Initiative (GENFI)

## Abstract

**Background:** Executive dysfunction is a core feature of frontotemporal dementia (FTD). Whilst there has been extensive research into such impairments in sporadic FTD, there has been little research in the familial forms.

**Methods:** 752 individuals were recruited in total: 214 *C9orf72*, 205 *GRN* and 86 *MAPT* mutation carriers, stratified into asymptomatic, prodromal and fully symptomatic, and 247 mutation negative controls. Attention and executive function were measured using the Weschler Memory Scale-Revised (WMS-R) Digit Span Backwards (DSB), the Wechsler Adult Intelligence Scale-Revised Digit Symbol task, the Trail Making Test Parts A and B, the Delis-Kaplan Executive Function System Color Word Interference Test and verbal fluency tasks (letter and category). Linear regression models with bootstrapping were used to assess differences between groups. Correlation of task score with disease severity was also performed, as well an analysis of the neuroanatomical correlates of each task.

**Results:** Fully symptomatic *C9orf72, GRN* and *MAPT* mutation carriers were significantly impaired on all tasks compared with controls (all p<0.001), except on the WMS-R DSB in the *MAPT* mutation carriers (p=0.147). Whilst asymptomatic and prodromal *C9orf72* individuals also demonstrated deficits compared with controls, neither the *GRN* or *MAPT* asymptomatic or prodromal mutation carriers showed significant differences. All tasks significantly correlated with disease severity in each of the genetic groups (all p<0.001).

**Conclusions:** Individuals with *C9orf72* mutations show difficulties with executive function from very early on in the disease and this continues to deteriorate with disease severity. In contrast, similar difficulties occur only in the later stages of the disease in *GRN* and *MAPT* mutation carriers. This differential performance across the genetic groups will be important in neuropsychological task selection in upcoming clinical trials.

## Introduction

Frontotemporal dementia (FTD) is a neurodegenerative disease that causes impairments in behaviour and cognition. Whilst a number of different changes in personality can occur such as apathy, loss of empathy and obsessive-compulsive behaviours (Warren et al., 2013), the core cognitive deficit is a change in executive function, a set of processes which includes inhibitory control, working memory and cognitive flexibility (Carlson et al., 2013).

Executive function has been extensively studied in sporadic FTD, where it has been demonstrated that such abilities are commonly compromised (Harciarek et al., 2013; Huey et al., 2009; Leslie et al., 2016; Moheb et al., 2017; Possin et al., 2013; Ramanan et al., 2017; Staffaroni et al., 2021; Tartaglia et al., 2012; van den Berg et al., 2017). However, there have been fewer studies examining changes in the familial forms of FTD due to mutations in chromosome 9 open reading frame 72 (*C9orf72*), progranulin (*GRN*) and microtubule associated protein tau (*MAPT*), which account for about one third of all FTD (Cheran et al., 2019; Hallam et al., 2014; Jiskoot et al., 2018; Lulé et al., 2020; Poos et al., 2020; Staffaroni et al., 2020).

The aim of this study was therefore to investigate executive function in a large cohort of presymptomatic and symptomatic individuals with familial FTD using participants from the GENetic Frontotemporal dementia Initiative (GENFI). In particular, we explore whether there are differences across the three main genetic causes and whether deficits occur in the presymptomatic stages of the disease. Such information will be important in guiding task selection for inclusion and outcome measures in upcoming therapeutic trials.

## Methods

### Cohort and protocol

The fifth GENFI data freeze included 831 individuals from 25 sites in Europe and Canada. 752 of these individuals had completed at least one of the executive function tasks in the GENFI neuropsychological battery: 214 carried the *C9orf72* expansion, 205 had a *GRN* mutation and 86 had a *MAPT* mutation; 247 individuals were mutation negative family members who acted as controls. Ethical approval was gained at each of the individual sites and all participants provided fully informed consent.

All participants underwent the standard GENFI protocol including the GENFI neuropsychological battery (Rohrer et al., 2015; Poos et al, 2022) as well as the Mini Mental State Examination (MMSE) and the CDR^®^ Dementia Staging Instrument with the National Alzheimer Coordinating Centre Frontotemporal Lobar Degeneration component (CDR^®^ plus NACC FTLD). The latter generates two types of scores: a sum of boxes (SB) score, and a global score which allows staging of the disease into 0, asymptomatic, 0.5, prodromal and 1 or more (1+), symptomatic (1 mild, 2 moderate and 3 severe). Symptomatic individuals were diagnosed according to current criteria (Rascovsky et al., 2011; Gorno-Tempini et al., 2011; Strong et al., 2017): 91 had bvFTD (*C9orf72* = 49, *GRN* = 24, *MAPT* = 18), 20 had primary progressive aphasia (*C9orf72* = 3, *GRN* = 16, *MAPT* = 1), and nine had FTD with amyotrophic lateral sclerosis (*C9orf72* = 9), whilst the other symptomatic participants consisted of smaller diagnostic groups including those with atypical parkinsonism. Demographic information can be found in Table.

Compared to controls, all three symptomatic groups were older (all p>0.001) and so too was the *GRN* prodromal group (p=0.004). The *MAPT* asymptomatic group were significantly younger than the controls (p=0.001). All three symptomatic groups were also significantly older than their asymptomatic (all p<0.001) and prodromal counterparts (all p<0.003). The *C9orf72* and *GRN* prodromal groups were also significantly older than their asymptomatic counterparts (p=0.025 and p=0.012 respectively). The *C9orf72* asymptomatic group was significantly older than the *MAPT* asymptomatic group (p=0.016) and the *GRN* symptomatic group wassignificantly older than the *MAPT* symptomatic group (p=0.040).

Differences in sex were found within the groups. *C9orf72* and *MAPT* symptomatic groups consisted of significantly more males than the control group (*X*^2^ (1) = 9.8, p=0.002; *X*^2^ (1) = 4.8, p=0.028 respectively) and there were more men in the *C9orf72* symptomatic group than the other *C9orf72* groups (asymptomatic: *X*^2^ (1) = 8.8, p=0.003; prodromal: *X*^2^ (1) = 6.4, p=0.012). This was also the case for the *MAPT* symptomatic carriers (asymptomatic: X^2^ (1) = 4.7, p=0.030; prodromal: *X*^2^ (1) = 5.1, p=0.023).

When investigating education across the groups, the *GRN* symptomatic individuals had significantly lower levels of education than the control group (p<0.001), the *GRN* asymptomatic group (p<0.01), and the *C9orf72* (p=0.035) and *MAPT* (p=0.030) symptomatic groups. The *C9orf72* symptomatic group had a lower level of education than the control group (p=0.010) and the *C9orf72* asymptomatic group (p=0.027).

### Executive function tasks

The following executive function tasks were included within the GENFI neuropsychology battery: the Wechsler Adult Intelligence Scale-Revised Digit Symbol substitution task (DSST) (Wechsler, 1981), the Weschler Memory Scale-Revised Digit Span Backwards (DSB) (Wechsler, 1997), the Delis-Kaplan Executive Function System (D-KEFS) Color-Word Interference Test (Color naming, Word naming and Ink color naming; Delis et al., 2001), and the Trail Making Test Parts A and B (TMT A and TMT B).

### Magnetic resonance image acquisition

T1-weighted magnetic resonance imaging (MRI) volumetric brain scans were performed on 703 participants as per the GENFI protocol (Rohrer et al., 2015). 55 images were removed as they either failed the quality control check for motion and scanner artefacts, or an abnormal finding was found in the form of significant vascular disease or other structural brain lesions. Subsequently, 648 scans were included in the analysis: 217 controls, 184 *C9orf72* expansion carriers, 172 *GRN* mutation carriers, and 75 *MAPT* mutation carriers.

### Statistical analysis

#### Healthy controls

To explore the normative performance on the tasks in the control group, percentile scores and cumulative frequencies were calculated for each, and a lower 5^th^ percentile was generated to indicate an abnormal score. t tests were performed to assess any differences on each of the tasks that were normally distributed, and Mann Whitney U tests for those that were not. Correlations between task performance and both age and education were calculated using Pearson’s correlation for normally distributed data and Spearman’s rank correlation for those that were not. Linear regressions were carried out to assess the impact of language (i.e. the language spoken by the participant) on each of the tasks within the control group. Pairwise post hoc comparisons were carried out to assess the difference between the groups if the overall model was significant. For data that was not normally distributed, bootstrapping with 2000 replications was used. All analyses were performed using Stata/IC (version 14.2).

#### Mutation carriers

Multiple linear regressions were carried out to assess performance on the tasks in each of the groups. Age, sex, education, and language were included in the models as covariates. Post hoc pairwise comparisons were calculated to assess the differences between the groups. For data that was not normal, bootstrapping with 2000 replications was used and the 95% bootstrapped confidence intervals reported.

A Pearson’s correlational analysis was conducted on each of the tasks to measure the association with disease severity and tasks performance using the CDR^®^ plus NACC FTLD SB score. For data that was not normally distributed, Spearman’s rank correlation (rho) was used instead.

To assess the impact of phenotype in the symptomatic groups, participants were grouped into bvFTD, PPA and FTD-ALS. Other phenotypes were not included in the analysis due to their low numbers. A linear regression was performed and included age, sex, and education as covariates in the model. For data that was not normally distributed, the model was bootstrapped (2000 replications).

#### Structural brain imaging analysis

An automated atlas segmentation propagation and label fusion strategy – (Geodesic Information Flow: GIF) (Cardoso et al., 2015) was used on the T1-weighted volumetric MRI scans to generate brain volumes of regions of interest known to be involved in executive function (Carlson et al., 2013; Cash et al., 2018): the orbitofrontal cortex (OFC), the dorsolateral prefrontal cortex (DLPFC), the ventromedial prefrontal cortex (VMPFC), the parietal lobe and the striatum. All the individual regional volumes were expressed as a percentage of total intracranial volume, as computed with SPM12 (Statistical Parametric Mapping, Welcome Trust Centre for Neuroimaging, London, UK) running under Matlab R2014b (Mathworks, USA: Malone et al., 2015). Using RStudio (version 1.2.1335, 2009-2019), partial correlations were performed to investigate the association between the brain regions and score on each of the executive function tasks, whilst taking into consideration disease severity as measured using the CDR^®^ plus NACC FTLD SB score, as well as the age of the participant.

## Results

### Healthy controls

#### Age

Increasing age correlated with worse performance on the DSST (r = -0.4, p<0.001), D-KEFS: Color (Rho = 0.3, p<0.001); D-KEFS: Word (Rho = 0.1, p=0.049); D-KEFS: Ink (Rho = 0.3, p<0.001), TMT A (Rho = 0.4, p<0.001) and TMT B (Rho = 0.3, p<0.001) but not the DSB. Performance across each decade can be found in Table S1.

#### Sex

There was a significant effect of sex on the DSST (T = 3.9, p<0.001, with females scoring higher) and TMT B (U = -2.1, p=0.035, with females performing quicker). No other differences were found between males and females on any of the other tasks. (Table S2).

#### Education

Across all of the tasks, there was a significant influence of education on task performance in the control group, with higher levels of education associated with better task performance (DSST: r = 0.4, p<0.001; DSB: r = 0.3, p<0.001; D-KEFS: Color: Rho = -0.2, p=0.004, D-KEFS: Word: Rho = -0.2, p=0.004; D-KEFS: Ink: Rho = -0.2, p<0.001; TMT A: Rho = -0.1, p=0.028; TMT B: Rho = -0.3, p< 0.001).

#### Language

Only the D-KEFS: Word and Ink tasks saw an overall influence of language on performance (D-KEFS: Word: Chi^2^(7) = 20.2, p=0.005; D-KEFS: Ink: Chi^2^(7) = 15.4, p=0.030, r^2^ = 0.056) (Table S3).

#### Percentile scores

Normative percentile scores were calculated on each of the tasks using the control data (Table S). A score below 38 on the DSST, below 3 on the DSB task would be considered abnormal (<5^th^ percentile). If it took more than 40 seconds, 31 seconds, and 71 seconds to complete the D-KEFS: Color, Word and Ink tasks respectively, and more than 48 seconds and 125 seconds for the TMT A and B tasks respectively, the participant’s performance would also be considered abnormal (<5^th^ percentile).

### Mutation carriers

#### Group comparisons

The means and standard deviations for the scores on the executive function tasks in each of the mutation carrier groups can be found in Table and Figure 1. The differences between the groups are shown in Tables S5 to 11 and Figure S.

**Figure 1:**
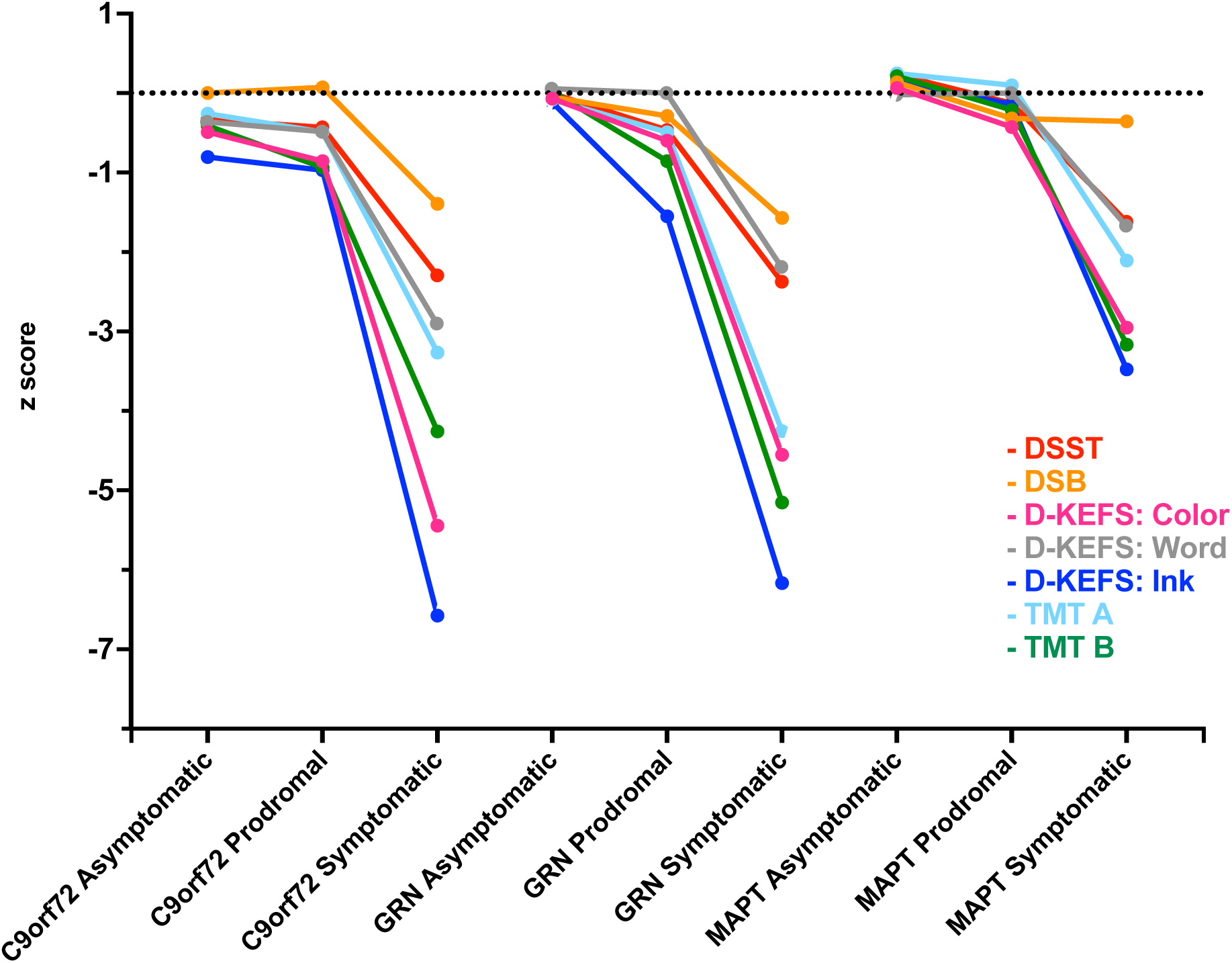
Performance of the mutation carrier groups on each of the executive function tasks expressed as a z-score to allow comparison across tasks. Abbreviations: DSST: Wechsler Adult Intelligence Scale-Revised Digit Symbol substitution task; DSB: Weschler Memory Scale-Revised Digit Span Backwards; D-KEFS: Delis Kaplan Executive Function System (Color-Word Inference Test – Color = Color naming, Word = Word naming, Ink = Ink color naming); TMT: Trail Making Test (A = Part A; B = Part B).

When compared to controls, all three symptomatic mutation carrier groups were significantly impaired on all executive function tasks compared to controls, as well as when compared to their asymptomatic and prodromal genetic groups, except for on the DSB task where no differences were seen between the different *MAPT* groups.

The *C9orf72* prodromal group were significantly impaired in comparison to controls on the D-KEFS: Color and Ink tasks and the TMT B with a trend towards a poorer performance on the DSST (p=0.066) and TMT A (p=0.099). No differences were seen between the prodromal *GRN* or *MAPT* groups and controls on any of the tasks.

The *C9orf72* asymptomatic group was significantly impaired compared to controls on all tasks except for the DSB and D-KEFS: Word tasks. No differences were seen between the asymptomatic *GRN* or *MAPT* groups and controls on any of the tasks.

When comparing between the genetic groups at each disease severity stage: for symptomatic mutation carriers, the *C9orf72* group performed significantly worse than the *MAPT* group on the DSB, the DSST, and the D-KEFS: Color and Ink tasks whilst the *GRN* symptomatic group performed worse than the *MAPT* symptomatic carriers on the DSB, DSST, and TMT A tasks; for prodromal mutation carriers, the *C9orf72* group performed significantly worse than the *MAPT* group on the D-KEFS: Ink task; for asymptomatic mutation carriers the *C9orf72* group performed significantly worse than the other two groups on the DSST, D-KEFS: Ink and TMT B tasks as well as worse than the *GRN* group on the D-KEFS: Color and TMT A tasks.

#### Correlation with disease severity

All tasks significantly correlated with disease severity as measured using the CDR^®^ plus NACC FTLD -SB score in each of the genetic groups (Table 1).

**Table 1:**
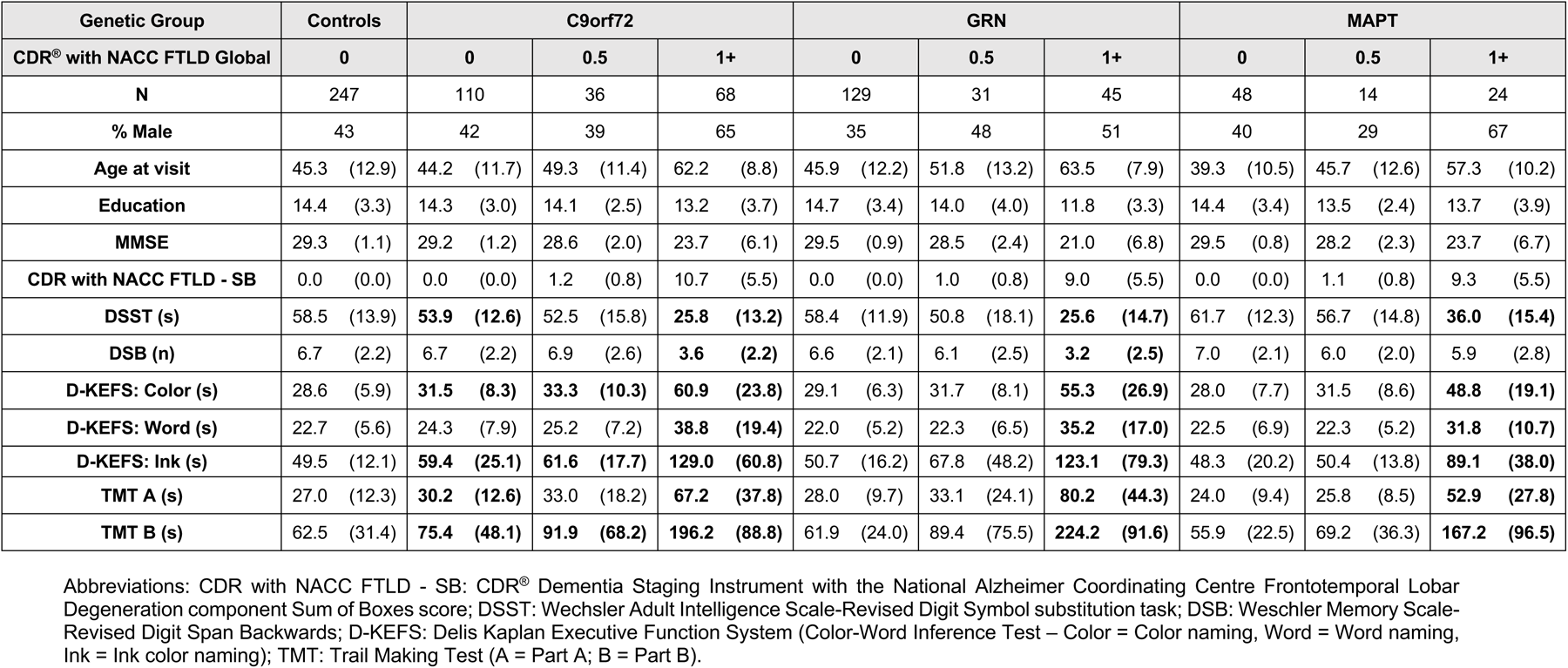
Demographic information and task performance for the participants split by genetic group and CDR^®^ plus NACC FTLD global score. Means and standard deviations (in parentheses) are given. Results in bold show significant differences between the mutation carrier groups and controls.

**Table 1:**
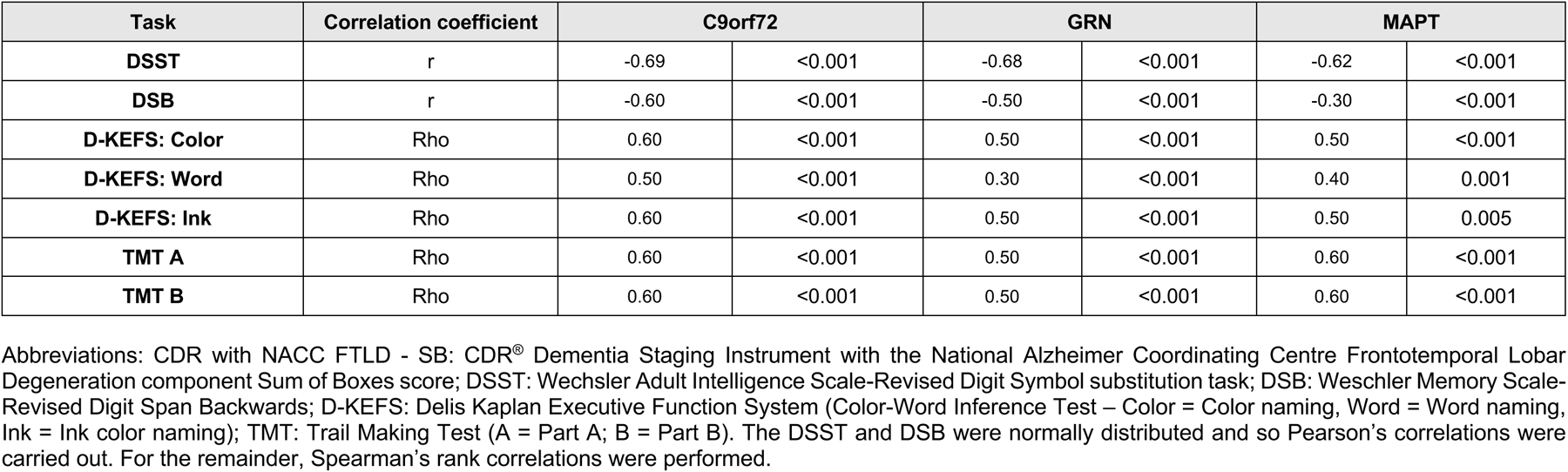
Correlations of each of the tasks with disease severity as measured using the CDR plus NACC FTLD - SB (for the three mutation carrier groups).

#### Phenotypic analysis

When compared to controls, all phenotypic groups were significantly impaired on all tasks of executive function (all p<0.001 except for the FTD-ALS group on the TMT A where p=0.016). The PPA group scored significantly worse on the DSB task than the bvFTD group (adjusted mean difference (AMD) = -1.5, p=0.011) and FTD-ALS groups (AMD = -1.5, p=0.012), and significantly worse than the bvFTD group on the TMT B (bvFTD: AMD = 56.2, p=0.011).

#### Imaging analysis (Table S12)

For *C9orf72* expansion carriers, correlations between task performance and regional brain volumes were seen with the dorsolateral prefrontal cortex (DSST, DSB and D-KEFS: Ink) and parietal cortex (DSST and TMT B), and also with the striatum on the DSST. For *GRN* mutation carriers, fewer significant correlations were seen: right orbitofrontal cortex with TMTB, and dorsolateral prefrontal cortex with D-KEFS: Color and Ink tasks. For *MAPT* mutation carriers, significant correlations were seen mainly with the dorsolateral prefrontal cortex (D-KEFS: Color and Ink and TMT B), and the striatum (DSST and D-KEFS: Color) as well as the left orbitfrontal cortex with D-KEFS: Ink).

## Discussion

In this study, the executive function abilities of a large cohort of individuals with genetic FTD were comprehensively assessed. It demonstrates that executive dysfunction is present in both individuals who are symptomatic of familial FTD across all three genetic groups, as well as in those individuals with a *C9orf72* expansion who are asymptomatic and prodromal. Neither the *GRN* nor the *MAPT* asymptomatic or prodromal mutation carriers showed significant differences on the executive function tasks compared to the control group suggesting that executive function changes occur later in the disorder than in *C9orf72*-associated FTD.

The control group data indicates that of all the demographic covariates, age and education were most associated with executive function score. This was to be expected as many studies have found that older age and lower education leads to greater impairment on many of the executive function tasks (Choi et al., 2014; Gaertner et al., 2018; Mathuranath et al., 2003; Tombaugh, 2004; Van der Elst et al., 2006). Only two tasks showed an effect of sex on score which is supported by previous work, with females performing better on the DSST, whilst males achieved better scores on the TMT B task (Kertzman et al., 2006; Majeres, 1983; Scheuringer et al., 2017). Finally, language did also have an influence on task performance, affecting those tests that had an element of language to them: the D-KEFS: Word and Ink tasks. This may well be due to the slight variation in the length of words in the different languages used in the study, which may take longer to pronounce. Further to note, is the differences in sample sizes across the control groups which may also have influenced the findings in this study.

As expected, and in line with the previous literature, the symptomatic individuals displayed executive dysfunction irrespective of genetic group (Cheran et al., 2019; Hallam et al., 2014; Jiskoot et al., 2018; Lulé et al., 2020; Poos et al., 2020; Staffaroni, Bajorek, et al., 2020). This was the case when they were compared to both the control group and their asymptomatic and prodromal counterparts. The only exception to this, was on the DSB task where the symptomatic *MAPT* mutation carriers were not significantly impaired.

The asymptomatic and prodromal *C9orf72* groups displayed early executive dysfunction in tasks assessing inhibition, set switching/cognitive flexibility, processing speed and general cognitive function, whilst working memory abilities were less affected early on. This is in line with a recent study that produced a cognitive composite for each of the genetic mutations that displayed widespread cognitive dysfunction in *C9orf72* mutation carriers, including deficits in executive function (Poos et al., 2022). Although two of the tasks in the prodromal group did not show a significant difference compared with controls despite this being seen in the asymptomatic group, it is likely that this was due to the smaller sample size in the prodromal group and thus, was not sufficiently powered to detect a deficit.

Neither the *GRN* nor the *MAPT* asymptomatic or prodromal carriers displayed any early executive dysfunction when compared to the controls. For the *GRN* mutation carriers, this is consistent with previous work that has demonstrated a rapid decline in symptoms in *GRN* mutation carriers during the first year of diagnosis compared to *C9orf72* and *MAPT* mutation carriers (Poos et al., 2020), with a rapid increase in atrophy rates after symptom onset (Staffaroni, Goh, et al., 2020), and the highest NfL levels of the three genetic groups in the symptomatic period, despite showing no difference to controls in the asymptomatic or prodromal period (Rojas et al., 2021). So, despite the clear deficit in executive function in the symptomatic phase of the disease, this work suggests that it is a symptom present later in the disease course and thus, may not be a useful marker of disease progression in primary or secondary prevention clinical trials. In contrast, it is likely that executive function is less affected in *MAPT* mutation carriers. Overall, the performance on the executive function tasks by the *MAPT* symptomatic mutation carriers was better than those of the other symptomatic genetic groups, with no significant difference being seen between the *MAPT* symptomatic group and the controls on the DSB task. The atrophy pattern in *MAPT* mutation carriers is much more localised with significant atrophy in the hippocampus, amygdala, and the temporal lobes (Bocchetta et al., 2021; Cash et al., 2018). These are regions usually associated with language and memory abilities of which impairments are present early on in *MAPT* mutation carriers (Moore et al., 2020; Pickering-Brown et al., 2008; Poos et al., 2022).

It was expected that individuals with a bvFTD diagnosis would perform worse than all other phenotypes on these executive tasks because it is a key diagnostic feature (Rascovsky et al., 2011) and there has been much evidence to support this in the sporadic literature (M. Harciarek & S. Cosentino, 2013; Huey et al., 2009; Leslie et al., 2016; Moheb et al., 2017; Possin et al., 2013; Ramanan et al., 2017; Staffaroni et al., 2021; Tartaglia et al., 2012; van den Berg et al., 2017). However, we find here that this was not the case on all tasks of executive function. Performance on the TMT B task was worst in those with a PPA diagnosis compared to those individuals with bvFTD, and performance on the DSB task was also worse in those with PPA compared to those with bvFTD or an ALS diagnosis. It is possible that this is because the DSB and TMT B tasks require some aspect of language function to be able to complete the test and thus, are not solely executive function tasks. The DSB task requires participants to access their lexicon to generate and produce number words, which is a particular problem if the individual is non-fluent. Furthermore, the TMT B task requires a knowledge of the alphabet. The human visual word store area or “letterbox” is found within the temporal lobe (Dehaene, 2013), a key region involved in PPA (Ruksenaite et al., 2021). It is therefore likely that executive function is not necessarily more impaired in PPA than it is in bvFTD and ALS, but rather the tasks are not solely executive tasks and so they are performing poorly on them due to the underlying language requirements. Despite this, all three phenotypes (bvFTD, PPA and ALS) displayed executive dysfunction when compared to controls which was expected and in line with prior work (Bettcher & Sturm, 2014; Michał Harciarek & Stephanie Cosentino, 2013; Taylor et al., 2013).

When looking at the mutation carriers as a whole, a clear decline in executive function is seen on all tasks for each genetic group as the disease progresses (when disease severity is (measured using the CDR plus the NACC-FTLD - SB score). This is consistent with previous work that demonstrates that function declines with disease severity in FTD (Nelson et al., 2021; Peakman et al., 2022).

The region of interest analysis revealed that in the *C9orf72* mutation carriers executive function score was associated with atrophy in the dorsolateral prefrontal cortex as well as the parietal lobe mainly. This is consistent with previous literature showing key involvement of the dorsolateral prefrontal cortex in executive function abilities (Dubreuil-Vall et al., 2019; Miller & Cohen, 2001; Ptak & Schnider, 2004), a region that is part of a wider fronto-parietal executive function network (Allen et al., 2011; Hartung et al., 2021; Hausman et al., 2022; Talwar et al., 2020). The DSST was associated with atrophy in multiple regions, likely due to the fact it assesses multiple cognitive processes including processing speed, working memory and reasoning (Rypma et al., 2006). In contrast to the *C9orf72* neural correlates, there were fewer correlates with the *GRN* mutation groups but consistent with involvement of the frontal lobe there were some associations with both the orbitofrontal and dorsolateral prefrontal cortices. Finally, when looking at the neural correlates of the *MAPT* mutation carriers, similar to the *GRN* mutation carriers, there is involvement of some frontal regions on a few of the tasks: orbitofrontal cortex on the D-KEFS: Ink task and the DLPFC on the TMT B and the D-KEFS: Color and Ink tasks. Striatal atrophy was also correlated with scores on the DSST and the D-KEFS: Color tasks, a region highly connected to the frontal lobe and well known to be associated with executive dysfunction when impaired (Faber et al., 2016; Tian et al., 2020).

Limitations to the study include the relatively small number of individuals investigated when breaking down the cohort into smaller groups. Further work is required to increase the sample size ensuring greater power and confidence in the results. A second limitation is the paucity of language-specific tasks available in the GENFI cognitive battery – future studies would examine the association of executive and language function across the different phenotypes, allowing us to break down the PPA group into the individual subgroups and understand further how language may be impacting performance, especially in the *GRN* mutation carriers.

To conclude, this study comprehensively assessed executive function abilities in a large cohort of individuals with genetic forms of FTD. It is clear that individuals with *C9orf72* mutations have difficulties with executive function from a very early stage in the disease and this continues to deteriorate with disease severity. In contrast, executive dysfunction occurs in the later stages of the disease in *GRN* and *MAPT* mutation carriers. Whilst it is assumed that executive dysfunction is a core feature of FTD, it appears that not all tasks measuring executive function do so equally across the genetic groups and so great care and consideration should be given when thinking about what tasks should be included as outcome measures in upcoming clinical trials based on the target genetic group and stage of the individuals being recruited.

## Data Availability

All data produced in the present study are available upon reasonable request to the authors

## Acknowledgements

We thank the research participants for their contribution to the study. The Dementia Research Centre is supported by Alzheimer’s Research UK, Alzheimer’s Society, Brain Research UK, and The Wolfson Foundation. This work was supported by the NIHR UCL/H Biomedical Research Centre, the Leonard Wolfson Experimental Neurology Centre (LWENC) Clinical Research Facility, and the UK Dementia Research Institute, which receives its funding from UK DRI Ltd, funded by the UK Medical Research Council, Alzheimer’s Society and Alzheimer’s Research UK. JDR is supported by the Miriam Marks Brain Research UK Senior Fellowship and has received funding from an MRC Clinician Scientist Fellowship (MR/M008525/1) and the NIHR Rare Disease Translational Research Collaboration (BRC149/NS/MH). This work was also supported by the MRC UK GENFI grant (MR/M023664/1), the Bluefield Project and the JPND GENFI-PROX grant (2019-02248). Several authors of this publication (JCvS, MS, RSV, AD, MO, RV, JDR) are members of the European Reference Network for Rare Neurological Diseases - Project ID No 739510. MB is supported by a Fellowship award from the Alzheimer’s Society, UK (AS-JF-19a-004-517). MB is also supported by the UK Dementia Research Institute which receives its funding from DRI Ltd, funded by the UK Medical Research Council, Alzheimer’s Society and Alzheimer’s Research UK. RC/CG are supported by a Frontotemporal Dementia Research Studentships in memory of David Blechner funded through The National Brain Appeal (RCN 290173). JBR is supported by the Wellcome Trust (103838), the Medical Research Council (MC_UU_00030/14; MR/T033371/1) and the NIHR Cambridge Biomedical Research Centre (NIHR203312). The views expressed are those of the authors and not necessarily those of the NIHR or the Department of Health and Social Care. The GIF template database includes volumetric MRI scans from the University College London Genetic FTD Initiative (GENFI) study (www.genfi.org.uk) which is funded by the Medical Research Council UK GENFI grant (MR/M023664/1). The GIF template database includes volumetric MRI scans from the University College London Genetic FTD Initiative (GENFI) study (www.genfi.org.uk) which is funded by the Medical Research Council UK GENFI grant (MR/M023664/1).

