## Supplemental Data for "Executive function deficits in genetic frontotemporal dementia: results from the GENFI study"

### Supplementary data

**Table S1: Relationship of age with score on each of the tasks in the control group. The groups are stratified by each of decade of life from 18 to 70+. The table shows the number of individuals in each group as well as the mean and standard deviation (SD) score on each task.**

| Group | Decade | DSST | | | DSB | | | D-KEFS: Color | | | D-KEFS: Word | | | D-KEFS: Ink | | | TMT A | | | TMT B | | |
|---|---|---|---|---|---|---|---|---|---|---|---|---|---|---|---|---|---|---|---|---|---|---|
| | | N | Mean | SD | N | Mean | SD | N | Mean | SD | N | Mean | SD | N | Mean | SD | N | Mean | SD | N | Mean | SD |
| Controls | 18-29.9 | 31 | 37.3 | 12.1 | 31 | 6.8 | 2.3 | 31 | 27.7 | 6.3 | 31 | 23.7 | 6.5 | 31 | 46.6 | 12.8 | 31 | 26.2 | 12.6 | 31 | 59.8 | 30.5 |
| | 30-39.9 | 64 | 43.3 | 15.1 | 64 | 6.9 | 2.1 | 64 | 26.7 | 4.7 | 64 | 20.9 | 3.7 | 64 | 46.2 | 10.0 | 64 | 21.9 | 6.5 | 64 | 51.7 | 15.0 |
| | 40-49.9 | 66 | 40.7 | 13.8 | 67 | 6.6 | 2.0 | 64 | 28.8 | 6.4 | 64 | 22.7 | 5.5 | 64 | 49.7 | 11.2 | 65 | 23.8 | 7.6 | 66 | 61.1 | 39.2 |
| | 50-59.9 | 45 | 42.9 | 10.2 | 45 | 6.9 | 2.4 | 45 | 29.2 | 6.3 | 45 | 23.0 | 7.6 | 45 | 50.9 | 13.2 | 45 | 28.1 | 11.6 | 43 | 61.8 | 26.6 |
| | 60-69.9 | 32 | 40.2 | 14.7 | 32 | 6.4 | 2.0 | 32 | 31.0 | 5.6 | 32 | 24.3 | 4.2 | 32 | 54.8 | 12.7 | 32 | 36.6 | 14.3 | 32 | 83.5 | 31.3 |
| | 70+ | 5 | 38.6 | 7.7 | 8 | 6.3 | 2.8 | 6 | 32.7 | 3.5 | 6 | 25.3 | 4.3 | 6 | 61.4 | 11.9 | 8 | 52.0 | 21.3 | 5 | 108.6 | 20.5 |

Abbreviations: DSST: Wechsler Adult Intelligence Scale-Revised Digit Symbol substitution task; DSB: Weschler Memory Scale-Revised Digit Span Backwards; D-KEFS: Delis Kaplan Executive Function System (Color-Word Inference Test – Color = Color naming, Word = Word naming, Ink = Ink color naming); TMT: Trail Making Test (A = Part A; B = Part B).

**Table S2: Relationship of sex with score on each of the tasks in the control group. Mean and standard deviations (SD) for the control group on each of the individual tasks when split into males and females. Tests in which the data were normally distributed were analysed using t tests were run, whilst for those that were not normally distributed Mann Whitney U tests were used.**

| Task | Females | | Males | | Significant difference between groups | |
|---|---|---|---|---|---|---|
| | Mean | SD | Mean | SD | T/U | P |
| **DSST** | 61.4 | 13.1 | 54.6 | 14.2 | 3.9 | <0.001 |
| **DSB** | 6.9 | 2.3 | 6.5 | 2.0 | 1.3 | 0.197 |
| **D-KEFS: Color** | 28.4 | 6.0 | 29.0 | 5.8 | -1.3 | 0.186 |
| **D-KEFS: Word** | 22.8 | 6.4 | 22.6 | 4.3 | -0.5 | 0.596 |
| **D-KEFS: Ink** | 48.7 | 12.1 | 50.7 | 12.0 | -1.4 | 0.177 |
| **TMT A** | 26.2 | 10.7 | 28.0 | 14.2 | -0.1 | 0.680 |
| **TMT B** | 57.8 | 22.5 | 68.9 | 39.6 | -2.1 | 0.035 |

Abbreviations: DSST: Wechsler Adult Intelligence Scale-Revised Digit Symbol substitution task; DSB: Weschler Memory Scale-Revised Digit Span Backwards; D-KEFS: Delis Kaplan Executive Function System (Color-Word Inference Test – Color = Color naming, Word = Word naming, Ink = Ink color naming); TMT: Trail Making Test (A = Part A; B = Part B).

**Table S3: Number of control individuals who performed the executive function tasks in each language, with the means and standard deviation (SD) for each group.**

| Language | DSST | | | DSB | | | D-KEFS: Color | | | D-KEFS: Word | | | D-KEFS: Ink | | | TMT A | | | TMT B | | |
|---|---|---|---|---|---|---|---|---|---|---|---|---|---|---|---|---|---|---|---|---|---|
| | N | Mean | SD | N | Mean | SD | N | Mean | SD | N | Mean | SD | N | Mean | SD | N | Mean | SD | N | Mean | SD |
| **English** | 52 | 59.2 | 13.4 | 52 | 7.3 | 2.5 | 52.0 | 28.1 | 6.5 | 52 | 24.0 | 7.4 | 52 | 51.4 | 12.7 | 52 | 26.0 | 15.9 | 51 | 57.6 | 28.3 |
| **Italian** | 31 | 54.0 | 16.4 | 33 | 6.1 | 1.5 | 30.0 | 28.9 | 5.6 | 30 | 22.2 | 7.1 | 30 | 53.4 | 12.2 | 32 | 28.2 | 14.2 | 31 | 57.2 | 17.9 |
| **Dutch** | 75 | 56.0 | 10.9 | 77 | 6.7 | 2.1 | 75.0 | 28.7 | 6.8 | 75 | 23.4 | 5.0 | 75 | 45.9 | 11.9 | 76 | 28.4 | 10.6 | 76 | 67.7 | 41.2 |
| **Swedish** | 12 | 59.3 | 16.7 | 12 | 5.9 | 1.9 | 12.0 | 31.6 | 6.3 | 12 | 23.3 | 3.2 | 12 | 54.3 | 11.7 | 12 | 24.9 | 10.5 | 12 | 73.2 | 38.4 |
| **French** | 27 | 58.4 | 14.2 | 27 | 6.9 | 2.2 | 27.0 | 27.4 | 3.3 | 27 | 21.2 | 3.2 | 27 | 50.5 | 10.8 | 27 | 23.6 | 6.4 | 25 | 54.0 | 14.3 |
| **Spanish** | 31 | 65.4 | 14.7 | 31 | 6.7 | 2.0 | 31.0 | 28.6 | 4.8 | 31 | 20.3 | 4.0 | 31 | 49.0 | 10.9 | 31 | 24.5 | 7.8 | 31 | 60.3 | 22.7 |
| **German** | 4 | 62.8 | 19.3 | 4 | 5.3 | 1.5 | 4.0 | 31.4 | 4.6 | 4 | 24.9 | 4.2 | 4 | 52.5 | 3.8 | 4 | 32.0 | 25.0 | 4 | 58.5 | 28.0 |
| **Portuguese** | 11 | 63.2 | 14.7 | 11 | 7.0 | 2.7 | 11.0 | 27.4 | 5.5 | 11 | 22.3 | 3.7 | 11 | 48.1 | 14.8 | 11 | 33.9 | 13.8 | 11 | 80.0 | 31.0 |

Abbreviations: DSST: Wechsler Adult Intelligence Scale-Revised Digit Symbol substitution task; DSB: Weschler Memory Scale-Revised Digit Span Backwards; D-KEFS: Delis Kaplan Executive Function System (Color-Word Inference Test – Color = Color naming, Word = Word naming, Ink = Ink color naming); TMT: Trail Making Test (A = Part A; B = Part B).

**Table S4: Percentile scores for each of the executive function tasks.**

| Percentile | DSST | DSB | DKEFS: Color (s) | DKEFS: Word (s) | DKEFS: Ink (s) | TMTA (s) | TMTB (s) |
|---|---|---|---|---|---|---|---|
| **5%** | 38 | 3 | 40 | 31 | 71 | 48 | 125 |
| **10%** | 42 | 4 | 37 | 28 | 65 | 39 | 92 |
| **25%** | 49 | 5 | 32 | 25 | 56 | 32 | 70 |
| **50%** | 59 | 7 | 28 | 22 | 49 | 24 | 56 |
| **75%** | 68 | 8 | 25 | 19 | 40 | 19 | 44 |
| **90%** | 76 | 10 | 22 | 17 | 35 | 16 | 36 |
| **95%** | 79 | 11 | 20 | 16 | 32 | 14 | 33 |

Abbreviations: DSST: Wechsler Adult Intelligence Scale-Revised Digit Symbol substitution task; DSB: Weschler Memory Scale-Revised Digit Span Backwards; D-KEFS: Delis Kaplan Executive Function System (Color-Word Inference Test – Color = Color naming, Word = Word naming, Ink = Ink color naming); TMT: Trail Making Test (A = Part A; B = Part B).

**Table S5: Wechsler Adult Intelligence Scale-Revised Digit Symbol substitution task: adjusted mean differences between the groups with p-values and 95% bootstrapped confidence intervals.**

| | | Control | C9orf72 | | | GRN | | | MAPT | | |
|---|---|---|---|---|---|---|---|---|---|---|---|
| | | | 0 | 0.5 | 1+ | 0 | 0.5 | 1+ | 0 | 0.5 | 1+ |
| Control | | | -4.81<br><0.001<br>-7.39 -2.23 | -3.81<br>0.066<br>-7.87 0.25 | -21.85<br><0.001<br>-25.44 -18.26 | -0.45<br>0.726<br>-2.95 2.06 | -3.65<br>0.102<br>-8.03 0.72 | -21.56<br><0.001<br>-25.59 -17.53 | 0.35<br>0.850<br>-3.26 3.95 | -1.85<br>0.555<br>-8.00 4.30 | -15.67<br><0.001<br>-20.89 -10.44 |
| C9orf72 | 0 | | | 1.00<br>0.652<br>-3.35 5.35 | -17.04<br><0.001<br>-20.96 -13.11 | 4.36<br>0.004<br>1.38 7.35 | 1.16<br>0.628<br>-3.53 5.84 | -16.75<br><0.001<br>-21.11 -12.40 | 5.16<br>0.011<br>1.20 9.12 | 2.96<br>0.361<br>-3.39 9.32 | -10.86<br><0.001<br>-16.34 -5.37 |
| | 0.5 | | | | -18.04<br><0.001<br>-22.89 -13.19 | 3.36<br>0.128<br>-0.97 7.69 | 0.00<br>0.956<br>-5.43 5.74 | -17.75<br><0.001<br>-23.00 -12.51 | 4.16<br>0.110<br>-0.94 9.26 | 1.96<br>0.585<br>-5.08 9.00 | -11.86<br><0.001<br>-18.07 -5.64 |
| | 1+ | | | | | 21.40<br><0.001<br>17.52 25.29 | 18.20<br><0.001<br>13.10 23.29 | 0.29<br>0.900<br>-4.24 4.82 | 22.20<br><0.001<br>17.37 27.03 | 20.00<br><0.001<br>13.23 26.77 | 6.18<br>0.032<br>0.52 11.85 |
| GRN | 0 | | | | | | -3.21<br>0.169<br>-7.78 1.37 | -21.11<br><0.001<br>-25.38 -16.84 | 0.79<br>0.694<br>-3.17 4.76 | -1.40<br>0.666<br>-7.77 4.97 | -15.22<br><0.001<br>-20.70 -9.74 |
| | 0.5 | | | | | | | -17.91<br><0.001<br>-23.30 -12.51 | 4.00<br>0.146<br>-1.40 9.40 | 1.81<br>0.627<br>-5.49 9.10 | -12.01<br><0.001<br>-18.42 -5.60 |
| | 1+ | | | | | | | | 21.91<br><0.001<br>16.73 27.09 | 19.71<br><0.001<br>12.68 26.74 | 5.90<br>0.055<br>-0.12 11.91 |
| MAPT | 0 | | | | | | | | | -2.20<br>0.528<br>-9.03 4.64 | -16.01<br><0.001<br>-22.11 -9.92 |
| | 0.5 | | | | | | | | | | -13.82<br><0.001<br>-21.56 -6.07 |
| | 1+ | | | | | | | | | | |

**Table S6: Weschler Memory Scale-Revised Digit Span Backwards: adjusted mean differences between the groups with p-values and 95% bootstrapped confidence intervals.**

| | | Control | C9orf72 | | | GRN | | | MAPT | | |
|---|---|---|---|---|---|---|---|---|---|---|---|
| | | | 0 | 0.5 | 1+ | 0 | 0.5 | 1+ | 0 | 0.5 | 1+ |
| Control | | | -0.05<br>0.846<br>-0.51 0.42 | 0.16<br>0.660<br>-0.56 0.89 | -2.35<br><0.001<br>-2.98 -1.72 | 0.03<br>0.888<br>-0.42 0.48 | -0.23<br>0.566<br>-1.01 0.55 | -2.44<br><0.001<br>-3.15 -1.73 | 0.04<br>0.894<br>-0.60 0.69 | -0.99<br>0.083<br>-2.10 0.13 | -0.66<br>0.147<br>-1.56 0.23 |
| C9orf72 | 0 | | | 0.21<br>0.599<br>-0.57 0.99 | -2.30<br><0.001<br>-3.00 -1.61 | 0.08<br>0.774<br>-0.46 0.61 | -0.18<br>0.669<br>-1.02 0.66 | -2.40<br><0.001<br>-3.17 -1.62 | 0.09<br>0.803<br>-0.62 0.80 | -0.94<br>0.110<br>-2.09 0.21 | -0.62<br>0.201<br>-1.56 0.33 |
| | 0.5 | | | | -2.51<br><0.001<br>-3.38 -1.65 | -0.13<br>0.739<br>-0.90 0.64 | 0.00<br>0.442<br>-1.39 0.61 | -2.60<br><0.001<br>-3.54 -1.67 | -0.12<br>0.798<br>-1.03 0.79 | -1.15<br>0.077<br>-2.42 0.12 | -0.82<br>0.134<br>-1.90 0.25 |
| | 1+ | | | | | 2.38<br><0.001<br>1.70 3.07 | 2.12<br><0.001<br>1.22 3.03 | -0.09<br>0.823<br>-0.89 0.70 | 2.40<br><0.001<br>1.54 3.25 | 1.36<br>0.028<br>0.14 2.58 | 1.69<br>0.001<br>0.72 2.66 |
| GRN | 0 | | | | | | -0.26<br>0.531<br>-1.08 0.56 | -2.47<br><0.001<br>-3.23 -1.72 | 0.01<br>0.973<br>-0.70 0.72 | -1.02<br>0.084<br>-2.17 0.14 | -0.69<br>0.148<br>-1.63 0.25 |
| | 0.5 | | | | | | | -2.21<br><0.001<br>-3.17 -1.26 | 0.27<br>0.579<br>-0.69 1.24 | -0.76<br>0.259<br>-2.08 0.56 | -0.43<br>0.447<br>-1.55 0.68 |
| | 1+ | | | | | | | | 2.49<br><0.001<br>1.57 3.40 | 1.46<br>0.024<br>0.19 2.72 | 1.78<br>0.001<br>0.75 2.81 |
| MAPT | 0 | | | | | | | | | -1.03<br>0.103<br>-2.27 0.21 | -0.71<br>0.191<br>-1.76 0.35 |
| | 0.5 | | | | | | | | | | 0.33<br>0.641<br>-1.04 1.70 |
| | 1+ | | | | | | | | | | |

**Table S7: Delis Kaplan Executive Function System Color-Word Inference Test – Color Naming: adjusted mean differences between the groups with p-values and 95% bootstrapped confidence intervals.**

| | | Control | C9ORF72 | | | GRN | | | MAPT | | |
|---|---|---|---|---|---|---|---|---|---|---|---|
| | | | 0 | 0.5 | 1+ | 0 | 0.5 | 1+ | 0 | 0.5 | 1+ |
| Control | | | 2.68<br>0.003<br>0.90 4.45 | 4.03<br>0.007<br>1.12 6.94 | 28.55<br><0.001<br>22.42 34.67 | 0.22<br>0.766<br>-1.24 1.68 | 1.82<br>0.154<br>-0.69 4.33 | 22.43<br><0.001<br>13.81 31.05 | 0.23<br>0.846<br>-2.10 2.56 | 2.70<br>0.166<br>-1.13 6.53 | 18.86<br><0.001<br>10.98 26.73 |
| C9ORF72 | 0 | | | 1.36<br>0.401<br>-1.81 4.52 | 25.87<br><0.001<br>19.56 32.18 | -2.45<br>0.017<br>-4.47 -0.43 | -0.85<br>0.567<br>-3.77 2.07 | 19.75<br><0.001<br>10.98 28.53 | -2.44<br>0.076<br>-5.14 0.26 | 0.03<br>0.990<br>-4.03 4.08 | 16.18<br><0.001<br>8.13 24.23 |
| | 0.5 | | | | 24.51<br><0.001<br>17.83 31.19 | -3.81<br>0.016<br>-6.90 -0.72 | 0.00<br>0.236<br>-5.87 1.45 | 18.40<br><0.001<br>9.41 27.38 | -3.80<br>0.040<br>-7.42 -0.18 | -1.33<br>0.566<br>-5.87 3.21 | 14.82<br><0.001<br>6.50 23.15 |
| | 1+ | | | | | -28.32<br><0.001<br>-34.53 -22.11 | -26.72<br><0.001<br>-33.02 -20.42 | -6.12<br>0.243<br>-16.39 4.16 | -28.31<br><0.001<br>-34.98 -21.65 | -25.84<br><0.001<br>-32.92 -18.77 | -9.69<br>0.043<br>-19.07 -0.31 |
| GRN | 0 | | | | | | 1.60<br>0.224<br>-0.98 4.18 | 22.21<br><0.001<br>13.51 30.90 | 0.01<br>0.994<br>-2.60 2.62 | 2.48<br>0.224<br>-1.52 6.48 | 18.63<br><0.001<br>10.63 26.64 |
| | 0.5 | | | | | | | 20.61<br><0.001<br>11.83 29.38 | -1.59<br>0.363<br>-5.02 1.84 | 0.88<br>0.694<br>-3.50 5.26 | 17.03<br><0.001<br>8.95 25.12 |
| | 1+ | | | | | | | | -22.20<br><0.001<br>-31.17 -13.22 | -19.73<br><0.001<br>-29.06 -10.40 | -3.57<br>0.544<br>-15.12 7.97 |
| MAPT | 0 | | | | | | | | | 2.47<br>0.266<br>-1.89 6.83 | 18.62<br><0.001<br>10.46 26.79 |
| | 0.5 | | | | | | | | | | 16.15<br><0.001<br>7.39 24.92 |
| | 1+ | | | | | | | | | | |

**Table S8: Delis Kaplan Executive Function System Color-Word Inference Test – Word Naming: adjusted mean differences between the groups with p-values and 95% bootstrapped confidence intervals.**

| | | Control | C9ORF72 | | | GRN | | | MAPT | | |
|---|---|---|---|---|---|---|---|---|---|---|---|
| | | | 0 | 0.5 | 1+ | 0 | 0.5 | 1+ | 0 | 0.5 | 1+ |
| Control | | | 1.38<br>0.106<br>-0.29 3.06 | 1.67<br>0.177<br>-0.75 4.09 | 14.41<br><0.001<br>9.06 19.75 | -0.62<br>0.317<br>-1.83 0.59 | -0.63<br>0.592<br>-2.92 1.67 | 10.56<br><0.001<br>5.37 15.75 | 0.10<br>0.932<br>-2.10 2.30 | -1.16<br>0.345<br>-3.58 1.25 | 8.68<br><0.001<br>3.89 13.46 |
| C9ORF72 | 0 | | | 0.29<br>0.837<br>-2.46 3.03 | 13.03<br><0.001<br>7.50 18.55 | -2.00<br>0.036<br>-3.87 -0.13 | -2.01<br>0.149<br>-4.75 0.72 | 9.17<br>0.001<br>3.69 14.66 | -1.29<br>0.320<br>-3.83 1.25 | -2.55<br>0.070<br>-5.30 0.21 | 7.29<br>0.005<br>2.23 12.35 |
| | 0.5 | | | | 12.74<br><0.001<br>7.16 18.32 | -2.29<br>0.078<br>-4.84 0.26 | 0.00<br>0.163<br>-5.53 0.93 | 8.89<br>0.002<br>3.21 14.56 | -1.58<br>0.313<br>-4.64 1.48 | -2.84<br>0.081<br>-6.02 0.35 | 7.00<br>0.010<br>1.67 12.34 |
| | 1+ | | | | | -15.03<br><0.001<br>-20.49 -9.56 | -15.04<br><0.001<br>-20.70 -9.38 | -3.85<br>0.306<br>-11.24 3.53 | -14.31<br><0.001<br>-20.09 -8.54 | -15.57<br><0.001<br>-21.35 -9.80 | -5.73<br>0.095<br>-12.47 1.01 |
| GRN | 0 | | | | | | -0.01<br>0.993<br>-2.30 2.28 | 11.17<br><0.001<br>6.08 16.27 | 0.71<br>0.535<br>-1.54 2.97 | -0.55<br>0.674<br>-3.10 2.01 | 9.29<br><0.001<br>4.43 14.16 |
| | 0.5 | | | | | | | 11.18<br><0.001<br>5.62 16.75 | 0.72<br>0.640<br>-2.31 3.76 | -0.54<br>0.739<br>-3.70 2.62 | 9.30<br><0.001<br>4.21 14.40 |
| | 1+ | | | | | | | | -10.46<br><0.001<br>-15.98 -4.94 | -11.72<br><0.001<br>-17.48 -5.96 | -1.88<br>0.603<br>-8.96 5.20 |
| MAPT | 0 | | | | | | | | | -1.26<br>0.429<br>-4.38 1.86 | 8.58<br>0.001<br>3.41 13.75 |
| | 0.5 | | | | | | | | | | 9.84<br><0.001<br>4.64 15.04 |
| | 1+ | | | | | | | | | | |

**Table S9: Delis Kaplan Executive Function System Color-Word Inference Test – Ink Naming: adjusted mean differences between the groups with p-values and 95% bootstrapped confidence intervals.**

| | | Control | C9orf72 | | | GRN | | | MAPT | | |
|---|---|---|---|---|---|---|---|---|---|---|---|
| | | | 0 | 0.5 | 1+ | 0 | 0.5 | 1+ | 0 | 0.5 | 1+ |
| Control | | | 10.19<br><0.001<br>5.58 14.79 | 10.31<br><0.001<br>4.88 15.74 | 64.33<br><0.001<br>48.61 80.05 | -0.11<br>0.948<br>-3.43 3.20 | 12.51<br>0.112<br>-2.93 27.94 | 57.87<br><0.001<br>27.60 88.13 | 2.02<br>0.499<br>-3.85 7.90 | 1.26<br>0.717<br>-5.55 8.06 | 32.32<br><0.001<br>17.89 46.74 |
| C9orf72 | 0 | | | 0.12<br>0.972<br>-6.76 7.01 | 54.15<br><0.001<br>37.71 70.58 | -10.30<br><0.001<br>-15.55 -5.04 | 2.32<br>0.776<br>-13.68 18.32 | 47.68<br>0.002<br>17.08 78.28 | -8.16<br>0.023<br>-15.19 -1.13 | -8.93<br>0.026<br>-16.79 -1.07 | 22.13<br>0.004<br>6.96 37.30 |
| | 0.5 | | | | 54.02<br><0.001<br>38.10 69.95 | -10.42<br>0.001<br>-16.42 -4.42 | 0.00<br>0.790<br>-13.98 18.38 | 47.56<br>0.002<br>17.34 77.77 | -8.28<br>0.035<br>-16.00 -0.57 | -9.05<br>0.032<br>-17.30 -0.80 | 22.01<br>0.004<br>7.03 36.98 |
| | 1+ | | | | | -64.44<br><0.001<br>-80.21 -48.68 | -51.83<br><0.001<br>-74.44 -29.21 | -6.47<br>0.705<br>-39.96 27.03 | -62.31<br><0.001<br>-79.26 -45.35 | -63.07<br><0.001<br>-80.25 -45.90 | -32.01<br>0.002<br>-52.51 -11.52 |
| GRN | 0 | | | | | | 12.62<br>0.107<br>-2.73 27.97 | 57.98<br><0.001<br>27.69 88.26 | 2.13<br>0.525<br>-4.44 8.71 | 1.37<br>0.723<br>-6.19 8.93 | 32.43<br><0.001<br>17.75 47.10 |
| | 0.5 | | | | | | | 45.36<br>0.010<br>10.65 80.07 | -10.48<br>0.211<br>-26.90 5.94 | -11.25<br>0.187<br>-27.95 5.46 | 19.81<br>0.073<br>-1.83 41.45 |
| | 1+ | | | | | | | | -55.84<br><0.001<br>-87.23 -24.45 | -56.61<br><0.001<br>-86.74 -26.48 | -25.55<br>0.132<br>-58.75 7.66 |
| MAPT | 0 | | | | | | | | | -0.77<br>0.864<br>-9.52 7.99 | 30.29<br><0.001<br>14.94 45.65 |
| | 0.5 | | | | | | | | | | 31.06<br><0.001<br>15.52 46.60 |
| | 1+ | | | | | | | | | | |

**Table S10: Trail Making Test Part A: adjusted mean differences between the groups with p-values and 95% bootstrapped confidence intervals.**

| | | Control | C9orf72 | | | GRN | | | MAPT | | |
|---|---|---|---|---|---|---|---|---|---|---|---|
| | | | 0 | 0.5 | 1+ | 0 | 0.5 | 1+ | 0 | 0.5 | 1+ |
| Control | | | 3.38<br>0.014<br>0.69 6.08 | 4.61<br>0.099<br>-0.87 10.08 | 30.48<br><0.001<br>21.51 39.44 | 0.25<br>0.835<br>-2.12 2.62 | 1.26<br>0.708<br>-5.35 7.88 | 40.56<br><0.001<br>28.24 52.88 | 0.09<br>0.955<br>-2.98 3.16 | -0.04<br>0.987<br>-4.41 4.34 | 20.38<br><0.001<br>10.50 30.26 |
| C9orf72 | 0 | | | 1.22<br>0.673<br>-4.46 6.91 | 27.09<br><0.001<br>18.13 36.06 | -3.13<br>0.037<br>-6.07 -0.19 | -2.12<br>0.537<br>-8.84 4.61 | 37.17<br><0.001<br>24.75 49.60 | -3.29<br>0.080<br>-6.98 0.39 | -3.42<br>0.150<br>-8.08 1.24 | 17.00<br>0.001<br>6.98 27.02 |
| | 0.5 | | | | 25.87<br><0.001<br>15.66 36.09 | -4.36<br>0.129<br>-9.98 1.27 | 0.00<br>0.432<br>-11.67 4.99 | 35.95<br><0.001<br>22.86 49.04 | -4.52<br>0.140<br>-10.51 1.48 | -4.64<br>0.158<br>-11.09 1.80 | 15.78<br>0.005<br>4.78 26.78 |
| | 1+ | | | | | -30.23<br><0.001<br>-39.14 -21.32 | -29.21<br><0.001<br>-40.28 -18.15 | 10.08<br>0.189<br>-4.95 25.11 | -30.39<br><0.001<br>-39.73 -21.05 | -30.52<br><0.001<br>-40.16 -20.87 | -10.09<br>0.123<br>-22.92 2.73 |
| GRN | 0 | | | | | | 1.01<br>0.761<br>-5.53 7.55 | 40.31<br><0.001<br>27.97 52.65 | -0.16<br>0.927<br>-3.62 3.30 | -0.29<br>0.902<br>-4.89 4.31 | 20.13<br><0.001<br>10.03 30.23 |
| | 0.5 | | | | | | | 39.29<br><0.001<br>25.19 53.39 | -1.17<br>0.750<br>-8.39 6.04 | -1.30<br>0.735<br>-8.85 6.25 | 19.12<br>0.002<br>7.31 30.93 |
| | 1+ | | | | | | | | -40.47<br><0.001<br>-53.06 -27.88 | -40.59<br><0.001<br>-53.39 -27.80 | -20.17<br>0.011<br>-35.81 -4.53 |
| MAPT | 0 | | | | | | | | | -0.13<br>0.960<br>-5.10 4.84 | 20.29<br><0.001<br>10.13 30.46 |
| | 0.5 | | | | | | | | | | 20.42<br><0.001<br>10.08 30.76 |
| | 1+ | | | | | | | | | | |

**Table S11: Trail Making Test Part B: adjusted mean differences between the groups with p-values and 95% bootstrapped confidence intervals.**

| | | Control | C9orf72 | | | GRN | | | MAPT | | |
|---|---|---|---|---|---|---|---|---|---|---|---|
| | | | 0 | 0.5 | 1+ | 0 | 0.5 | 1+ | 0 | 0.5 | 1+ |
| Control | | | 12.83<br>0.006<br>3.59 22.08 | 27.23<br>0.004<br>8.61 45.85 | 112.59<br><0.001<br>89.05 136.13 | -4.04<br>0.174<br>-9.87 1.78 | 17.79<br>0.119<br>-4.58 40.17 | 130.71<br><0.001<br>101.63 159.80 | 0.57<br>0.888<br>-7.32 8.46 | 10.86<br>0.214<br>-6.28 27.99 | 96.19<br><0.001<br>58.86 133.53 |
| C9orf72 | 0 | | | 14.40<br>0.160<br>-5.69 34.48 | 99.76<br><0.001<br>74.86 124.65 | -16.88<br>0.001<br>-26.72 -7.04 | 4.96<br>0.679<br>-18.57 28.49 | 117.88<br><0.001<br>87.34 148.42 | -12.26<br>0.024<br>-22.88 -1.65 | -1.98<br>0.837<br>-20.81 16.85 | 83.36<br><0.001<br>45.29 121.43 |
| | 0.5 | | | | 85.36<br><0.001<br>55.42 115.30 | -31.27<br>0.001<br>-50.23 -12.32 | 0.00<br>0.525<br>-38.51 19.64 | 103.48<br><0.001<br>68.36 138.61 | -26.66<br>0.006<br>-45.69 -7.63 | -16.37<br>0.188<br>-40.78 8.03 | 68.96<br>0.001<br>27.62 110.31 |
| | 1+ | | | | | -116.64<br><0.001<br>-140.08 -93.19 | -94.80<br><0.001<br>-127.19 -62.40 | 18.12<br>0.335<br>-18.72 54.96 | -112.02<br><0.001<br>-136.58 -87.46 | -101.74<br><0.001<br>-130.42 -73.05 | -16.40<br>0.451<br>-59.06 26.27 |
| GRN | 0 | | | | | | 21.84<br>0.056<br>-0.59 44.27 | 134.76<br><0.001<br>105.76 163.76 | 4.61<br>0.284<br>-3.82 13.05 | 14.90<br>0.100<br>-2.85 32.65 | 100.24<br><0.001<br>62.71 137.77 |
| | 0.5 | | | | | | | 112.92<br><0.001<br>74.83 151.01 | -17.23<br>0.145<br>-40.38 5.93 | -6.94<br>0.629<br>-35.06 21.18 | 78.40<br><0.001<br>34.42 122.38 |
| | 1+ | | | | | | | | -130.14<br><0.001<br>-160.21 -100.08 | -119.86<br><0.001<br>-154.14 -85.58 | -34.52<br>0.154<br>-81.99 12.95 |
| MAPT | 0 | | | | | | | | | 10.29<br>0.257<br>-7.50 28.07 | 95.63<br><0.001<br>57.75 133.51 |
| | 0.5 | | | | | | | | | | 85.34<br><0.001<br>45.40 125.28 |
| | 1+ | | | | | | | | | | |

**Table S12: Partial correlations between task score and regional braoin volume for each for a) *C9orf72*, b) GRN and c) MAPT mutation carriers.**

**a)**

| **Brain region** | | ***DSST*** | | ***DSB*** | | ***TMT A*** | | ***TMT B*** | | ***D-KEFS: Color*** | | ***D-KEFS: Word*** | | ***D-KEFS: Ink*** | |
|---|---|---|---|---|---|---|---|---|---|---|---|---|---|---|---|
| | | ***Rho*** | ***p*** | ***Rho*** | ***p*** | ***Rho*** | ***p*** | ***Rho*** | ***p*** | ***Rho*** | ***p*** | ***Rho*** | ***p*** | ***Rho*** | ***p*** |
| Orbitofrontal cortex | Left | 0.08 | 0.275 | 0.07 | 0.309 | 0.08 | 0.310 | 0.02 | 0.787 | 0.03 | 0.711 | -0.02 | 0.752 | -0.01 | 0.859 |
| | Right | **0.18** | **0.012** | 0.07 | 0.320 | -0.03 | 0.722 | -0.06 | 0.408 | -0.02 | 0.768 | 0.02 | 0.795 | 0.01 | 0.912 |
| Dorsolateral prefrontal cortex | Left | **0.25** | **<0.001** | **0.14** | **0.047** | -0.11 | 0.141 | -0.08 | 0.305 | -0.08 | 0.299 | -0.06 | 0.388 | **-0.20** | **0.007** |
| | Right | **0.22** | **0.002** | **0.15** | **0.043** | -0.05 | 0.542 | -0.05 | 0.525 | -0.08 | 0.260 | 0.02 | 0.805 | **-0.21** | **0.005** |
| Ventromedial prefrontal cortex | Left | **0.17** | **0.020** | 0.13 | 0.063 | -0.07 | 0.325 | -0.05 | 0.537 | -0.03 | 0.637 | -0.07 | 0.368 | 0.05 | 0.532 |
| | Right | **0.16** | **0.034** | 0.10 | 0.150 | -0.07 | 0.364 | -0.04 | 0.555 | -0.04 | 0.634 | 0.02 | 0.822 | 0.03 | 0.695 |
| Parietal cortex | Left | **0.22** | **0.002** | 0.10 | 0.174 | -0.11 | 0.134 | **-0.16** | **0.030** | 0.01 | 0.896 | -0.07 | 0.380 | -0.14 | 0.058 |
| | Right | **0.28** | **0.000** | 0.09 | 0.222 | -0.14 | 0.059 | **-0.16** | **0.033** | 0.00 | 0.998 | -0.01 | 0.925 | -0.09 | 0.257 |
| Striatum | Left | **0.15** | **0.036** | 0.08 | 0.296 | -0.07 | 0.379 | -0.05 | 0.480 | -0.09 | 0.216 | -0.09 | 0.235 | -0.03 | 0.659 |
| | Right | 0.13 | 0.086 | 0.02 | 0.734 | -0.03 | 0.688 | 0.01 | 0.884 | -0.09 | 0.247 | -0.03 | 0.670 | 0.00 | 0.961 |

**b)**

| **Brain region** | | ***DSST*** | | ***DSB*** | | ***TMT A*** | | ***TMT B*** | | ***D-KEFS: Color*** | | ***D-KEFS: Word*** | | ***D-KEFS: Ink*** | |
|---|---|---|---|---|---|---|---|---|---|---|---|---|---|---|---|
| | | ***Rho*** | ***p*** | ***Rho*** | ***p*** | ***Rho*** | ***p*** | ***Rho*** | ***p*** | ***Rho*** | ***p*** | ***Rho*** | ***p*** | ***Rho*** | ***p*** |
| Orbitofrontal cortex | Left | -0.01 | 0.909 | 0.05 | 0.523 | -0.06 | 0.431 | -0.05 | 0.487 | -0.04 | 0.564 | -0.02 | 0.752 | 0.00 | 0.952 |
| | Right | 0.00 | 0.992 | -0.03 | 0.723 | 0.08 | 0.285 | **0.15** | **0.050** | 0.11 | 0.125 | 0.02 | 0.795 | 0.12 | 0.121 |
| Dorsolateral prefrontal cortex | Left | 0.11 | 0.137 | 0.13 | 0.073 | -0.14 | 0.055 | -0.08 | 0.268 | **-0.20** | **0.008** | -0.06 | 0.388 | -0.14 | 0.061 |
| | Right | 0.08 | 0.276 | 0.03 | 0.662 | -0.08 | 0.275 | -0.07 | 0.330 | **-0.21** | **0.005** | 0.02 | 0.805 | **-0.19** | **0.013** |
| Ventromedial prefrontal cortex | Left | -0.07 | 0.321 | -0.03 | 0.643 | 0.04 | 0.543 | 0.12 | 0.126 | 0.04 | 0.605 | -0.07 | 0.368 | 0.07 | 0.392 |
| | Right | -0.06 | 0.451 | 0.02 | 0.828 | 0.08 | 0.277 | 0.11 | 0.151 | -0.03 | 0.710 | 0.02 | 0.822 | 0.03 | 0.682 |
| Parietal cortex | Left | 0.07 | 0.325 | 0.04 | 0.617 | -0.05 | 0.459 | -0.09 | 0.243 | -0.07 | 0.373 | -0.07 | 0.380 | 0.03 | 0.700 |
| | Right | 0.02 | 0.833 | 0.01 | 0.915 | 0.02 | 0.734 | 0.00 | 0.996 | 0.03 | 0.649 | -0.01 | 0.925 | 0.10 | 0.190 |
| Striatum | Left | -0.02 | 0.825 | 0.09 | 0.212 | 0.01 | 0.907 | 0.01 | 0.879 | -0.12 | 0.100 | -0.09 | 0.235 | 0.06 | 0.451 |
| | Right | -0.06 | 0.401 | 0.07 | 0.331 | 0.03 | 0.665 | 0.06 | 0.458 | -0.08 | 0.262 | -0.03 | 0.670 | 0.09 | 0.215 |

**c)**

| Brain region | | *DSST* | | *DSB* | | *TMT A* | | *TMT B* | | *D-KEFS: Color* | | *D-KEFS: Word* | | *D-KEFS: Ink* | |
|---|---|---|---|---|---|---|---|---|---|---|---|---|---|---|---|
| | | *Rho* | *p* | *Rho* | *p* | *Rho* | *p* | *Rho* | *p* | *Rho* | *p* | *Rho* | *p* | *Rho* | *p* |
| Orbitofrontal cortex | Left | -0.06 | 0.617 | -0.15 | 0.190 | 0.09 | 0.473 | -0.06 | 0.640 | -0.04 | 0.763 | 0.13 | 0.295 | **-0.24** | **0.045** |
| | Right | 0.09 | 0.450 | -0.09 | 0.436 | 0.05 | 0.690 | -0.22 | 0.065 | 0.20 | 0.091 | 0.19 | 0.106 | -0.09 | 0.451 |
| Dorsolateral prefrontal cortex | Left | 0.13 | 0.281 | -0.16 | 0.187 | -0.12 | 0.299 | **-0.27** | **0.020** | -0.06 | 0.622 | -0.04 | 0.720 | -0.18 | 0.124 |
| | Right | 0.20 | 0.094 | -0.12 | 0.304 | -0.13 | 0.288 | **-0.27** | **0.019** | **-0.32** | **0.006** | -0.21 | 0.081 | **-0.34** | **0.003** |
| Ventromedial prefrontal cortex | Left | 0.12 | 0.307 | 0.01 | 0.936 | -0.02 | 0.892 | -0.12 | 0.305 | 0.04 | 0.721 | -0.07 | 0.548 | -0.06 | 0.618 |
| | Right | 0.03 | 0.824 | -0.04 | 0.759 | 0.05 | 0.700 | -0.03 | 0.804 | 0.03 | 0.783 | -0.04 | 0.745 | 0.03 | 0.790 |
| Parietal cortex | Left | 0.02 | 0.861 | -0.17 | 0.153 | 0.15 | 0.204 | 0.07 | 0.550 | -0.02 | 0.878 | 0.04 | 0.737 | -0.06 | 0.626 |
| | Right | 0.09 | 0.463 | -0.05 | 0.668 | 0.12 | 0.326 | 0.04 | 0.706 | 0.10 | 0.390 | 0.08 | 0.516 | 0.00 | 0.987 |
| Striatum | Left | **0.35** | **0.002** | 0.01 | 0.932 | -0.12 | 0.293 | -0.22 | 0.065 | **-0.26** | **0.028** | -0.17 | 0.153 | -0.20 | 0.087 |
| | Right | **0.35** | **0.003** | 0.02 | 0.875 | -0.14 | 0.240 | -0.23 | 0.050 | -0.22 | 0.061 | -0.16 | 0.178 | -0.20 | 0.088 |

Abbreviations: DSST: Wechsler Adult Intelligence Scale-Revised Digit Symbol substitution task; DSB: Weschler Memory Scale-Revised Digit Span Backwards; D-KEFS: Delis Kaplan Executive Function System (Color-Word Inference Test – Color = Color naming, Word = Word naming, Ink = Ink color naming); TMT: Trail Making Test (A = Part A; B = Part B).

***Figure S1:*** **Scores on each of the executive function tasks across all groups. *indicates a significant difference between the groups at p<0.05. Differences are only shown between each group and controls, and between mutation carrier groups within a specific mutation. Differences between groups across different mutation groups are not displayed. DSST: Wechsler Adult Intelligence Scale-Revised Digit Symbol substitution task; DSB: Weschler Memory Scale-Revised Digit Span Backwards; D-KEFS: Delis Kaplan Executive Function System (Color-Word Inference Test – Color = Color naming, Word = Word naming, Ink = Ink color naming); TMT: Trail Making Test (A = Part A; B = Part B).**

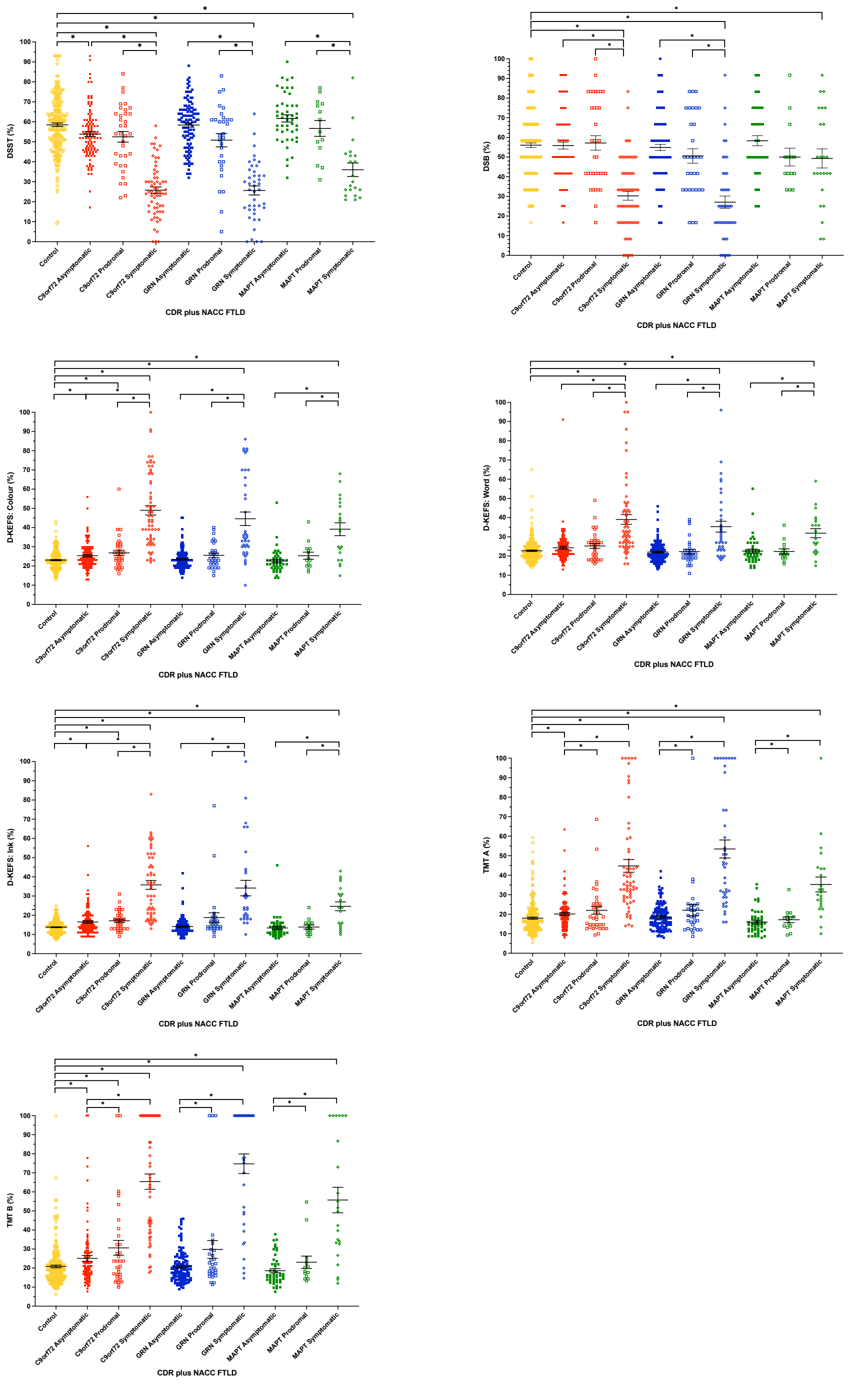

### Appendix

*Co-author list:*

Aitana Sogorb Esteve PhD[1,2], Martina Bocchetta PhD[1], David L Thomas PhD[3], Henrik Zetterberg PhD[2,4], Imogen J Swift MSc[5,2], Jennifer Nicholas PhD[6], Kiran Samra MRCP[5], Rachelle Shafei MRCP[1], Carolyn Timberlake[7], Thomas Cope MRCP PhD[8], Timothy Rittman MRCP PhD[9], Antonella Alberici MD[10], Enrico Premi MD[11], Roberto Gasparotti MD[12], Emanuele Buratti PhD[13,14], Valentina Cantoni MSc[15], Andrea Arighi MD[16], Chiara Fenoglio PhD[17], Elio Scarpini MD[17,18], Maria Serpente PhD[18], Stefano Floro MD[18], Vittoria Borracci[18], Giacomina Rossi[19], Giorgio Giaccone[19], Giuseppe Di Fede MD[19], Paola Caroppo MD PhD[19], Pietro Tiraboschi[19], Sara Prioni[19], Veronica Redaelli[19], David Tang-Wai[20], Ekaterina Rogaeva[21], Miguel Castelo-Branco MD PhD[22], Morris Freedman[23], Ron Keren[24], Sandra Black[25], Sara Mitchell[26], Christen Shoesmith[27], Robart Bartha PhD[28], Rosa Rademakers PhD[29], Jackie Poos MSc[30], Janne M Papma[31], Lucia Giannini MD[31], Rick van Minkelen[32], Benedetta Nacmias[33,34], Camilla Ferrari[33], Cristina Polito[35], Gemma Lombardi[36], Valentina Bessi[33], Michele Veldsman[37], Abbe Ullgren[38], Linn Öijerstedt[39,40], Vesna Jelic[41], Paul Thompson PhD[42], Albert Lladó MD, PhD[43], Anna Antonell PhD[44], Jordi Juncà-Parella MD, PhD[44], Mircea Balasa MD PhD[44], Nuria Bargalló MD PhD[45], Sergi Borrego-Ecija[46], Jordi Sarto[44], Ana Verdelho MD PhD[47], Carolina Maruta PhD[48], Catarina B Ferreira[49], Gabriel Miltenberger MD PhD[50], Frederico Simões do[51], Alazne Gabilondo MD[52,53], Jorge Villanua MD PhD[54], Marta Cañada[55], Mikel Tainta MD[56], Myriam Barandiaran PhD[52,53], Patricia Alves PhD[56,57], Benjamin Bender[58], Carlo Wilke[59,60], Lisa Graf[59], Annick Vogels[61], Mathieu Vandenbulcke[62], Philip Van Damme[63], Rose Bruffaerts MD PhD[64,65], Koen Poesen MD PhD[66], Pedro Rosa-Neto[67], Serge Gauthier[68], Agnès Camuzat[69], Daisy Rinaldi[70,71,69], Sabrina Sayah[72], Catharina Prix[73], Elisabeth Wlasich[74], Olivia Wagemann[75], Sandra Loosli[74], Sonja Schönecker[74], Tobias Hoegen[74], Jolina Lombardi[76], Sarah Anderl-Straub[76], Marijne Vandebergh PhD[77,78], Ilse Dewachter PhD[79], Adeline Rollin[80], Gregory Kuchcinski[81,80,82], Maxime Bertoux PhD[83,80], Thibaud Lebouvier[81,80,82], Vincent Deramecourt[84,80,82], Sanna Hannonen MD[85], Juhana Hakumaki[85], Miguel Tábuas-Pereira MD[86,22,87], Maria João Leitão MSc[88], Maria Rosario Almeida PhD[89], João Durães MD[86,90,91], Marisa Lima MSc[92,93], Miguel Castelo-Branco Md, PhD[94,95,96], João Lemos MD, PhD[97,95,93]

### *Co-author affiliations:*

[1]Department of Neurodegenerative Disease, Dementia Research Centre, UCL Queen Square Institute of Neurology, London, UK, [2]UK Dementia Research Institute at University College London, UCL Queen Square Institute of Neurology, London, UK, [3]Neuroimaging Analysis Centre, Department of Brain Repair and Rehabilitation, UCL Queen Square Institute of Neurology, London, UK, [4]Department

of Psychiatry and Neurochemistry, The Sahlgrenska Academy at the University of Gothenburg, UCL Queen Square Institute of Neurology, Mölndal, Sweden, [5]Department of Neurodegenerative Disease, Dementia Research Centre, London, UK, [6]Department of Medical Statistics, London School of Hygiene and Tropical Medicine, UCL Queen Square Institute of Neurology, London, UK, [7]Department of Clinical Neurosciences, UCL Queen Square Institute of Neurology, Cambridge, UK, [8]Department of Clinical Neuroscience, University of Cambridge, Cambridge, UK, [9]Department of Clinical Neurosciences, University of Cambridge, Cambridge, UK, [10]Centre for Neurodegenerative Disorders, ASST Brescia Hospital, University of Cambridge, Brescia, Italy, [11]Stroke Unit, ASST Brescia Hospital, University of Brescia, Brescia, Italy, [12]Neuroradiology Unit, University of Brescia, Brescia, Italy, [13]Molecular Pathology Laboratory, International Centre for Genetic Engineering and Biotechnology (ICGEB), 34149 Trieste, Trieste, Italy, [14]Brescia, Italy, [15]Centre for Neurodegenerative Disorders, Department of Clinical and Experimental Sciences, Brescia, Italy, [16]Fondazione IRCCS Ca' Granda Ospedale Maggiore Policlinico, Neurodegenerative Diseases Unit, University of Brescia, Milan, Italy, [17]University of Milan, Centro Dino Ferrari, Milan, Italy, [18]Fondazione IRCCS Ca' Granda Ospedale Maggiore Policlinico, Neurodegenerative Diseases Unit, Milan, Italy, [19]Fondazione IRCCS Istituto Neurologico Carlo Besta, Milano, Italy, [20]The University Health Network, Krembil Research Institute, Toronto, Canada, [21]Tanz Centre for Research in Neurodegenerative Diseases, University of Toronto, Toronto, Canada, [22]Faculty of Medicine, University of Coimbra, Coimbra, Portugal, [23]Baycrest Health Sciences, Rotman Research Institute, Toronto, Canada, [24]The University Health Network, Toronto Rehabilitation Institute, University of Toronto, Toronto, Canada, [25]Sunnybrook Health Sciences Centre, Sunnybrook Research Institute, Toronto, Canada, [26]Sunnybrook Health Sciences Centre, Sunnybrook Research Institute, University of Toronto, Toronto, Canada, [27]Department of Clinical Neurological Sciences, University of Western Ontario, University of Toronto, Ontario, Canada, [28]Department of Medical Biophysics, The University of Western Ontario, London, Ontario, Canada, [29]Center for Molecular Neurology, University of Antwerp, London, Belguim, [30]Department of Neurology, Erasmus Medical Center, Antwerp, Rotterdam, Netherlands, [31]Department of Neurology, Erasmus Medical Center, Rotterdam, Netherlands, [32]Department of Clinical Genetics, Erasmus Medical Center, Rotterdam, Netherlands, [33]Department of Neuroscience, Psychology, Drug Research and Child Health, University of Florence, Florence, Italy, [34]IRCCS Fondazione Don Carlo Gnocchi, Florence, Italy, [35]Department of Biomedical, Experimental and Clinical Sciences "Mario Serio", Florence, Italy, [36]IRCCS Fondazione Don Carlo Gnocchi, Nuclear Medicine Unit, Florence, Italy, [37]Nuffield Department of Clinical Neurosciences, Medical Sciences Division, Oxford, UK, [38]Center for Alzheimer Research, Division of Neurogeriatrics, University of Oxford, Stockholm, Sweden, [39]Center for Alzheimer Research, Division of Neurogeriatrics, Department of Neurobiology, Care Sciences and Society,

Karolinska Institutet, Bioclinicum, Solna, Sweden, [40]Unit for Hereditary Dementias, Theme Aging, Karolinska Institutet, Solna, Sweden, [41]Division of Clinical Geriatrics, Karolinska Institutet, Karolinska University Hospital, Sweden, [42]Division of Neuroscience and Experimental Psychology, Wolfson Molecular Imaging Centre, Stockholm, Manchester, UK, [43]Alzheimer's disease and Other Cognitive Disorders Unit, Neurology Service, University of Manchester, Barcelona, Spain, [44]Alzheimer's disease and Other Cognitive Disorders Unit, Neurology Service, Hospital Clínic, Barcelona, Spain, [45]Imaging Diagnostic Center, Hospital Clínic, Hospital Clínic, Barcelona, Spain, [46]Alzheimer's disease and Other Cognitive Disorders Unit, Neurology Service, Barcelona, Spain, [47]Department of Neurosciences and Mental Health, Centro Hospitalar Lisboa Norte - Hospital de Santa Maria & Faculty of Medicine, Hospital Clínic, Lisbon, Portugal, [48]Laboratory of Language Research, Centro de Estudos Egas Moniz, Faculty of Medicine, University of Lisbon, Lisbon, Portugal, [49]Laboratory of Neurosciences, Faculty of Medicine, University of Lisbon, Lisbon, Portugal, [50]Faculty of Medicine, University of Lisbon, University of Lisbon, Lisbon, Portugal, [51]Faculdade de Medicina, Universidade Católica Portuguesa, Lisbon, Portugal, [52]Cognitive Disorders Unit, Department of Neurology, San Sebastian, Gipuzkoa, Spain, [53]Neuroscience Area, Biodonostia Health Research Insitute, Donostia University Hospital, San Sebastian, Gipuzkoa, Spain, [54]OSATEK, University of Donostia, San Sebastian, Gipuzkoa, Spain, [55]CITA Alzheimer, San Sebastian, Gipuzkoa, Spain, [56]Neuroscience Area, Biodonostia Health Research Insitute, San Sebastian, Gipuzkoa, Spain, [57]Department of Educational Psychology and Psychobiology, Faculty of Education, Logroño, Spain, [58]Department of Diagnostic and Interventional Neuroradiology, University of Tübingen, International University of La Rioja, Tübingen, Germany, [59]Department of Neurodegenerative Diseases, Hertie-Institute for Clinical Brain Research and Center of Neurology, Tübingen, Germany, [60]Center for Neurodegenerative Diseases (DZNE), University of Tübingen, Tübingen, Germany, [61]Department of Human Genetics, KU Leuven, University of Tübingen, Leuven, Belgium, [62]Geriatric Psychiatry Service, University Hospitals Leuven, Leuven, Belgium, [63]Neurology Service, University Hospitals Leuven, Leuven, Belgium, [64]Department of Biomedical Sciences, University of Antwerp, Antwerp, Belgium, [65]Biomedical Research Institute, Hasselt University, 3500 Hasselt, Hasselt, Belgium, [66]Laboratory for Molecular Neurobiomarker Research, KU Leuven, Leuven, Belgium, [67]Translational Neuroimaging Laboratory, McGill Centre for Studies in Aging, Montreal, Québec, Canada, [68]Alzheimer Disease Research Unit, McGill Centre for Studies in Aging, Department of Neurology & Neurosurgery, McGill University, Montreal, Québec, Canada, [69]Sorbonne Université, Paris Brain Institute – Institut du Cerveau – ICM, Inserm U1127, CNRS UMR 7225, Paris, France, [70]Centre de référence des démences rares ou précoces, IM2A, Département de Neurologie, AP-HP - Hôpital Pitié-Salpêtrière, Paris, France, [71]Département de Neurologie, AP-HP - Hôpital Pitié-Salpêtrière (DMU Neurosciences Paris 6), AP-HP - Hôpital Pitié-Salpêtrière (DMU Neurosciences Paris 6), Paris,

France, [72]Sorbonne Université, Paris Brain Institute – Institut du Cerveau – ICM, Inserm U1127, CNRS UMR 7225, F-75013, Paris, France, [73]Neurologische Klinik, Ludwig-Maximilians-Universität München, AP-HP - Hôpital Pitié-Salpêtrière, Munich, Germany, [74]Neurologische Klinik, Ludwig-Maximilians-Universität München, Munich, Germany, [75](No affiliation data provided), [76]Department of Neurology, University of Ulm, Ulm, Germany, [77]VIB Center for Molecular Neurology, VIB, Antwerp, Belgium, [78]Department of Biomedical Sciences, University of Antwerp, Antwerp, Belgium, [79]Biomedical Research Institute, Hasselt University, Diepenbeek, Belgium, [80]CHU, CNR-MAJ, Lille, France, [81]Univ Lille, Labex Distalz, LiCEND, Lille, France, [82]Inserm 1172, Labex Distalz, LiCEND, Lille, France, [83]Inserm 1172, Lille, France, [84]Univ Lille, Lille, France, [85](No affiliation data provided), [86]Neurology Department, Centro Hospitalar e Universitario de Coimbra, Coimbra, Portugal, [87]Center for Innovative Biomedicine and Biotechnology (CIBB), University of Coimbra, Coimbra, Portugal, [88]Center for Innovative Biomedicine and Biotechnology (CIBB), Universidade de Coimbra, Coimbra, Portugal, [89]Centre of Neurosciences and Cell Biology, University of Coimbra, Coimbra, Portugal, [90]Faculty of Medicine, University of Coimbra, Portugal, Coimbra, Portugal, [91]Center for Innovative Biomedicine and Biotechnology (CIBB), University of Coimbra, Coimbra, Coimbra, Portugal, [92]Neurology Department, Centro Hospitalar e Universitário de Coimbra, Coimbra, Portugal, [93]Center for Innovative Biomedicine and Biotechnology (CIBB), University of Coimbra, Coimbra, [94]Institute of Nuclear Sciences Applied to Health (ICNAS), University of Coimbra, Coimbra, Portugal, [95]Faculty of Medicine, University of Coimbra, Coimbra, Portugal, [96]Institute for Biomedical Imaging and Life Sciences (CNC.IBILI), University of Coimbra, Coimbra, Portugal, [97]Neurology Department, Centro Hospitalar e Universitário de Coimbra, Coimbra, Portugal.
